## Supplemental Materials 1-5 for "Pharmaceutical Payments to Japanese Board-Certified Infectious Disease Specialists: A Four-year Retrospective Analysis of Payments from 92 Pharmaceutical Companies between 2016 and 2019"

Supplemental Material 1. Distribution of payment values per specialist

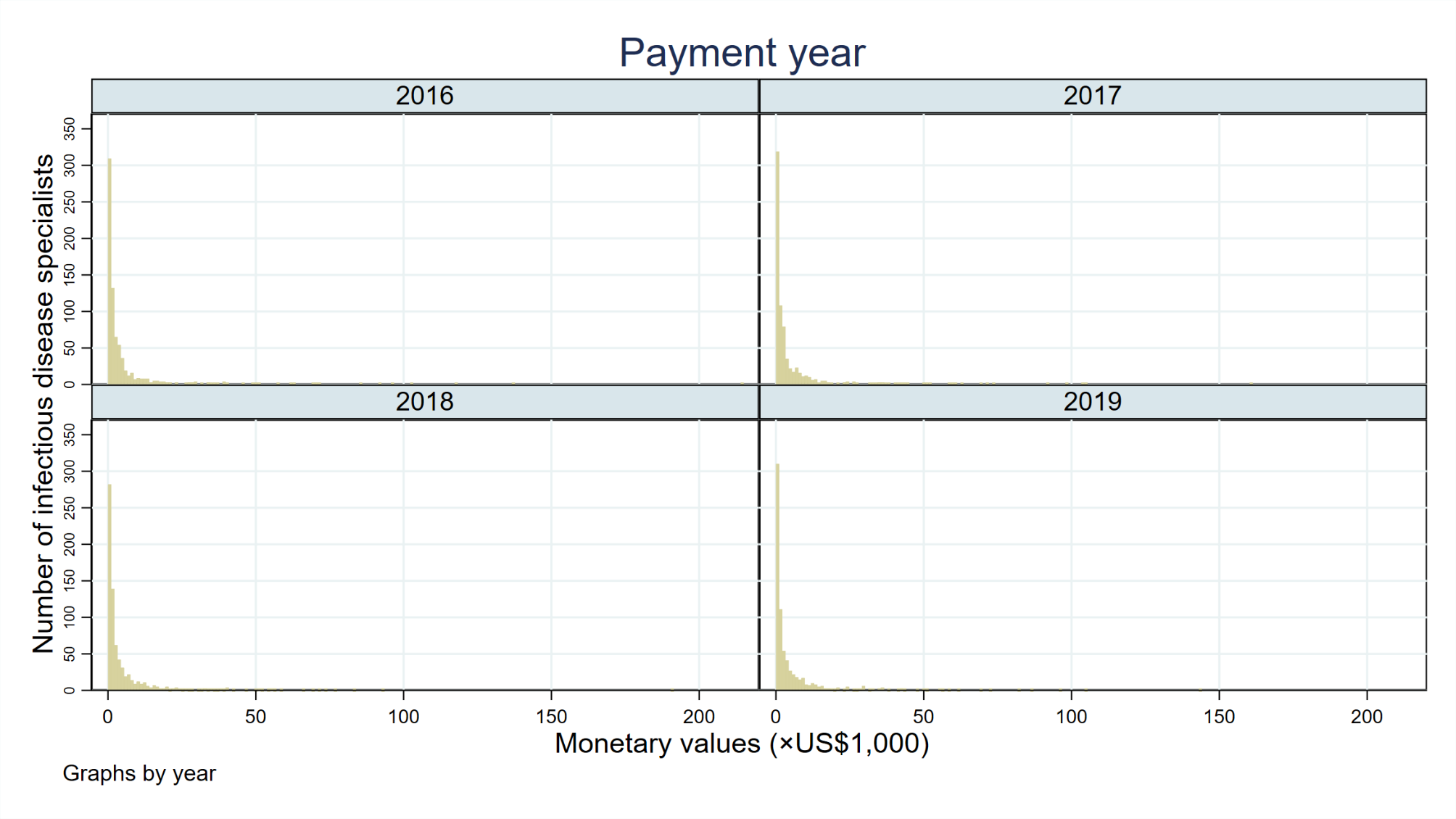

Supplemental Material 2. Payment concentration

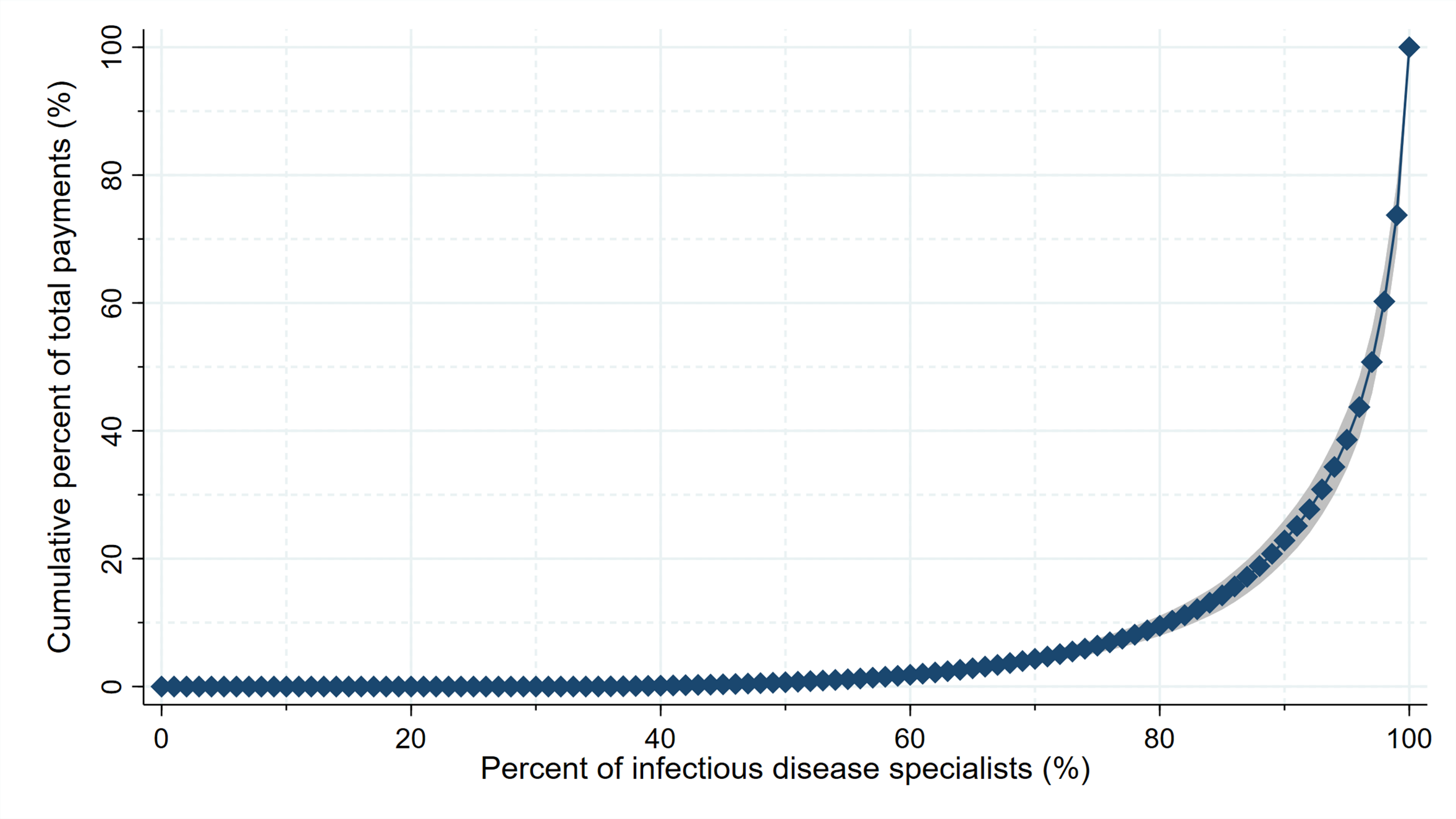

Supplemental Material 3. Payment category by company

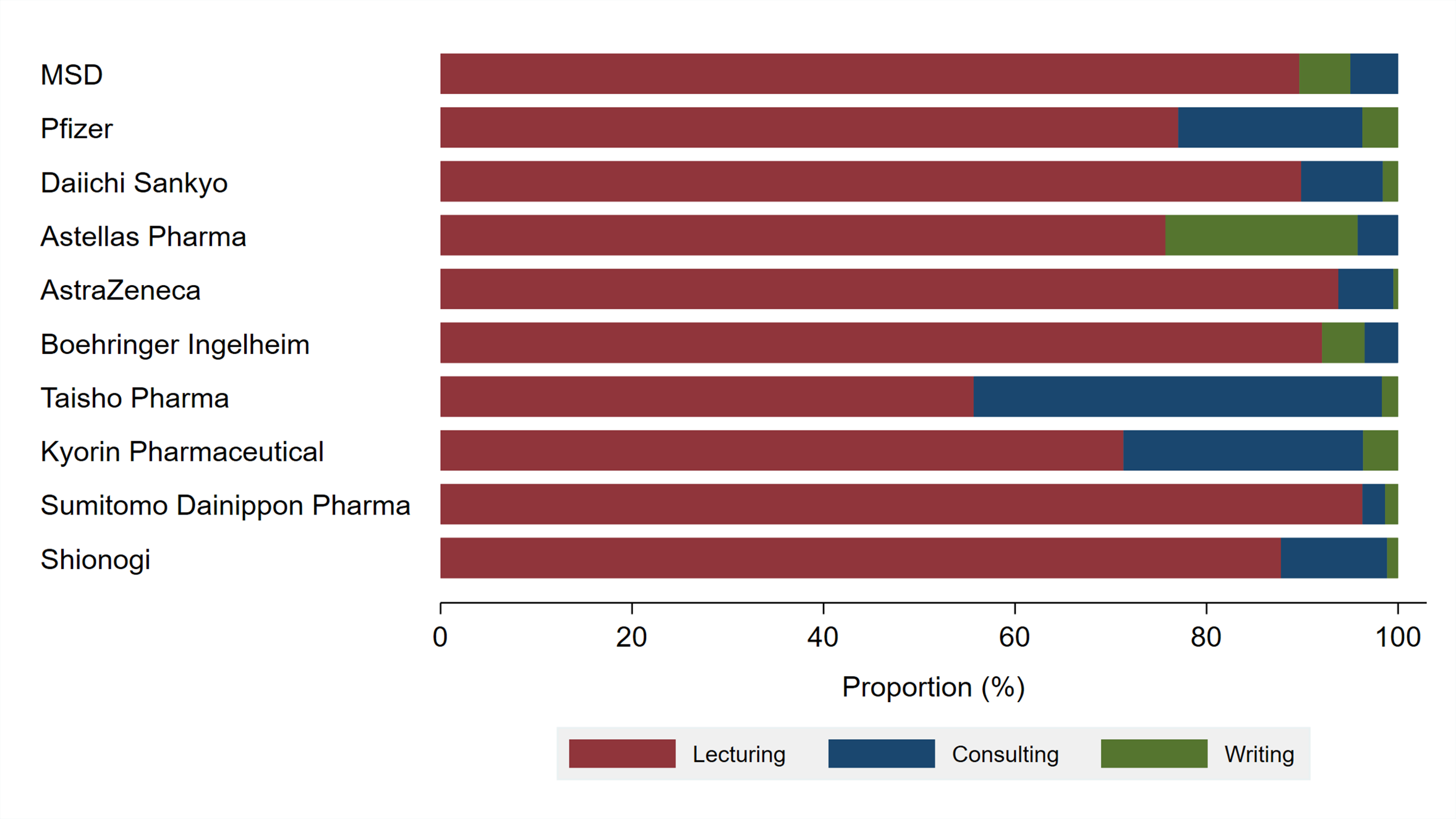

Supplemental Material 4. New and additional indications for infectious diseases in Japan between 2015 and 2019

| Brand name | Name | Pharmaceutical companies | Approval date | Price per drug unit, US$* | Indication | Category |
| --- | --- | --- | --- | --- | --- | --- |
| ZERBAXA | Ceftolozane sulfate/Tazobactam sodium | Manufacturer and distributor: MSD K. K | December 20, 2019 | $59 (1.5g/bottle) | Treatment of sepsis and pneumonia caused by Serratia and Hemophilus influenza | Additional indication |
| LASVIC | Lascufloxacin hydrochloride | Manufacturer and distributor: Kyorin Pharmaceuticals | September 20, 2019 | $3 (75mg/pill)  $37 (150mg/ intravenous infusion kit) | Treatment of laryngopharyngitis, tonsillitis, acute bronchitis, pneumonia, secondary infection of chronic respiratory disease, otitis media and sinusitis | New approval |
| MAVIRET | Glecaprevir hydrate/Pibrentasvir | Manufacturer and distributor: AbbVie GK | August 22, 2019 | $168/pill | Improvement of viremia in patients with chronic hepatitis C or compensated cirrhosis type C | Additional indication |
| AZIMYCIN | Azithromycin hydrate | Manufacturer and distributor: Senju Pharmaceuticals  Distributor: Takeda Pharmaceutical | June 18, 2019 | $3 (1%1ml eye-drops) | Treatment of conjunctivitis, blepharitis, hordeolum and dacryocystitis | New approval |
| SYMTUZA | Darunavir ethanolate/Cobicistat /Emtricitabine/Tenofovir alafenamide fumarate | Manufacturer and distributor: Janssen Pharmaceutical K. K | June 18, 2019 | $44/pill | Treatment of HIV-1 infection | New approval |
| INAVIR | Laninamivir octanoate hydrate | Manufacturer and distributor: Daiichi Sankyo | June 18, 2019 | $20 (20mg/inhalation kit)  $39 (160mg/bottle) | Treatment of influenza A or B virus infection | New approval |
| GENVOYA | Elvitegravir/Cobicistat /Emtricitabine/Tenofovir alafenamide fumarate | Manufacturer and distributor: Gilead Sciences | May 22, 2019 | $65/pill | Treatment of HIV-1 infection | Additional indication |
| BIKTARVY | Bictegravir sodium/ Emtricitabine/Tenofovir alafenamide fumarate | Manufacturer and distributor: Gilead Sciences | March 26, 2019 | $65/pill | Treatment of HIV-1 infection | New approval |
| ZERBAXA | Ceftolozane sulfate/Tazobactam sodium | Manufacturer and distributor: MSD K. K | January 8, 2019 | $59 (1.5g/bottle) | Treatment of cystitis, pyelonephritis, peritonitis, intra-abdominal abscess, cholecystitis and liver abscess | New approval |
| EPCLUSA | Sofosbuvir/Velpatasvir | Manufacturer and distributor: Gilead Sciences | January 8, 2019 | $562/pill | Improvement of viremia in patients with chronic hepatitis C or compensated cirrhosis type C who have previously been treated.  Improvement of viremia in patients with decompensated cirrhosis type C | New approval |
| REBETROL | Ribavirin | Manufacturer and distributor: ViiV Healthcare  Distributor: GlaxoSmithKline K. K  Promotional partner: Shionogi | January 8, 2019 | $4 (200mg) | Improvement of viremia in patients with chronic hepatitis C or compensated cirrhosis type C who have previously been treated | Additional indication |
| JULUCA | Dolutegravir sodium/Rilpivirine hydrochloride | Manufacturer and distributor: ViiV Healthcare  Distributor: GlaxoSmithKline K. K | November 26, 2018 | $50/pill | Treatment of HIV-1 infection | New approval |
| ODEFSEY | Rilpivirine hydrochloride /Emtricitabine/Tenofovir alafenamide fumarate | Manufacturer and distributor: Janssen Pharmaceutical K. K | August 21, 2018 | $56/pill | Treatment of HIV-1 infection | New approval |
| ISENTRESS | Raltegravir potassium | Manufacturer and distributor: MSD K. K | May 14, 2018 | $15 (400mg/pill)  $15 (600mg/pill) | Treatment of HIV infection | New approval |
| KAKETSUKEN | Emulsion-adjuvanted cell-culture derived influenza HA vaccine (H5N1) | Manufacturer and distributor: The Chemo-Sero-Therapeutic Research Institute | March 23, 2018 | NA | Prevention of pandemic influenza (H5N1) | Additional indication |
| KAKETSUKEN | Emulsion-adjuvanted cell-culture derived influenza HA vaccine (prototype) | Manufacturer and distributor: The Chemo-Sero-Therapeutic Research Institute | March 23, 2018 | NA | Prevention of pandemic influenza | Additional indication |
| XOFLUZA | Baloxavir marboxil | Manufacturer and distributor: Shionogi | February 23, 2018 | $14 (10mg/pill)  $22 (20mg/pill) | Treatment of influenza A or B virus infections | New approval |
| HARVONI | Ledipasvir acetonate/Sofosbuvir | Manufacturer and distributor: Gilead Sciences | February 16, 2018 | $509/pill | Improvement of viremia in patients with chronic hepatitis C or compensated cirrhosis type C in serogroup 2 | Additional indication |
| ZINPLAVA | Bezlotoxumab | Manufacturer and distributor: MSD K. K | September 27, 2017 | $3081 (625mg/25ml bottle) | Prevention of recurrent Clostridium difficile infection | New approval |
| MAVIRET | Glecaprevir hydrate/ Pibrentasvir | Manufacturer and distributor: AbbVie GK | September 27, 2017 | $168/pill | Improvement of viremia in patients with chronic hepatitis C or compensated cirrhosis type C | New approval |
| TAMIFLU | Oseltamivir phosphate | Manufacturer and distributor: Chugai Pharmaceutical | March 24, 2017 | $2 (75mg/pill)  $2 (3%/g dry syrup) | Treatment of influenza A or B virus infection | Additional indication |
| SOVALDI | Sofosbuvir | Manufacturer and distributor: Gilead Sciences | March 24, 2017 | $395 (400mg/pill) | Improvement of viremia in patients with chronic hepatitis C or compensated cirrhosis type C in neither Serogroup 1 (genotype 1) nor Serogroup 2 (genotype 2) | Additional indication |
| REBETOL | Ribavirin | Manufacturer and distributor: MSD K. K | March 24, 2017 | $4 (200mg/pill) | Improvement of viremia in patients with chronic hepatitis C or compensated cirrhosis type C in neither Serogroup 1 (genotype 1) nor Serogroup 2 (genotype 2) | Additional indication |
| OZEX | Tosufloxacin tosilate hydrate | Manufacturer and distributor: Fujifilm Toyama Chemical | March 2, 2017 | $1.1 (60mg/pill)  $0.5 (75mg/pill)  $0.6 (150mg/pill)  $1.2 (0.3%/ml eye-drops) | Treatment of mycoplasma pneumonia caused by Mycoplasma pneumoniae | Additional indication |
| RIAMET | Artemether/Lumefantrine | Manufacturer and distributor: Novartis Pharma K. K | December 19, 2016 | $2/pill | Treatment of malaria | New approval |
| VAXEM HIB | Hemophilus influenzae type b vaccine adsorbed | Manufacturer and distributor: Takeda Pharmaceutical | December 19, 2016 | NA | Prophylaxis of Hemophilus influenzae type b infections | Additional indication |
| XIMENCY | Daclatasvir hydrochloride/ Asunaprevir/Beclabuvir hydrochloride | Manufacturer and distributor: Bristol Myers Squibb | December 19, 2016 | $99/pill | Improvement of viremia in patients with chronic hepatitis C or compensated cirrhosis type C in serogroup 1 (genotype 1) | New approval |
| DESCOVY | Emtricitabine/ Tenofovir alafenamide fumarate | Manufacturer and distributor: Gilead Sciences | December 9, 2016 | $26/pill (LT)  $37/pill (HT) | Treatment of HIV-1 infection | New approval |
| PREZCOBIX | Darunavir ethanolate/ Cobicistat | Manufacturer and distributor: Janssen Pharmaceutical K. K | November 22, 2016 | $19 | Treatment of HIV infection | New approval |
| ERELSA | Elbasvir | Manufacturer and distributor: MSD K. K | September 28, 2016 | $223 (50mg/pill) | Improvement of viremia in patients with chronic hepatitis C or compensated cirrhosis type C in serogroup 1 (genotype 1) | New approval |
| VIEKIRAX | Ombitasvir hydrate/ Paritaprevir hydrate/ Ritonavir | Manufacturer and distributor: AbbVie GK | September 28, 2016 | $204/pill | Improvement of viremia in patients with chronic hepatitis C in serogroup 2 (genotype 2) | Additional indication |
| GRAZYNA | Grazoprevir hydrate | Manufacturer and distributor: MSD K. K | September 28, 2016 | $80 (50mg/pill) | Improvement of viremia in patients with chronic hepatitis C or compensated cirrhosis type C in serogroup 1 (genotype 1) | New approval |
| REBETOL | Ribavirin | Manufacturer and distributor: MSD K. K | September 28, 2016 | $4 (200mg/capsule) | Improvement of viremia in patients with chronic hepatitis C in serogroup 2 | Additional indication |
| INAVIR | Laninamivir octanoate hydrate | Manufacturer and distributor: Daiichi Sankyo | August 26, 2016 | $20 (20mg/inhalation kit)  $39 (160mg/bottle) | Prophylaxis of influenza A or B virus infections | Additional indication |
| GENVOYA | Elvitegravir/Cobicistat/ Emtricitabine/Tenofovir alafenamide fumarate | Manufacturer and distributor: Gilead Sciences | June 17, 2016 | $65/pill | Treatment of HIV-1 infection | New approval |
| MALARONE | Atovaquone/Proguanil hydrochloride | Manufacturer and distributor: GlaxoSmithKline K. K | March 28, 2016 | $5/pill (adult)  $2/pill (children) | Treatment and prevention of malaria | Additional indication |
| PRIMAQUINE | Primaquine phosphate | Manufacturer and distributor: Sanofi S.A. | March 28, 2016 | $21 (15mg/pill) | Treatment of malaria caused by Plasmodium vivax and P. ovale | New approval |
| BIKEN | Freeze-dried live attenuated varicella vaccine | Manufacturer and distributor: The Research Foundation for Microbial Diseases of Osaka University  Distributor: Mitsubishi Tanabe Pharma Corporation | March 18, 2016 | NA | Prevention of herpes zoster in individuals 50 years of age and older | Additional indication |
| KITASATO DAIICHI SANKYO | Adsorbed cell culture-derived influenza vaccine (H5N1) | Manufacturer and distributor: Daiichi Sankyo  Distributor: Kitasato Pharmaceutical Industry | March 18, 2016 | NA | Prevention of pandemic influenza (H5N1) | Additional indication |
| TRIBIK | Adsorbed diphtheria-purified pertussis-tetanus combined vaccine | Manufacturer and distributor: The Research Foundation for Microbial Diseases of Osaka University  Distributor: Mitsubishi Tanabe Pharma Corporation | February 29, 2016 | NA | Prevention of pertussis, diphtheria and tetanus | Additional indication |
| VAXEM HIB | Hemophilus influenzae type b vaccine absorbed | Manufacturer and distributor: Takeda Pharmaceutical | January 22, 2016 | NA | Prophylaxis of Hemophilus influenzae type b infections | New approval |
| REMICADE | Infliximab | Manufacturer and distributor: Mitsubishi Tanabe Pharma Corporation  Distributor: Janssen Biotech | December 21, 2015 | $648(100mg bottle) | Treatment of acute-phase Kawasaki's disease in patients who have not responded sufficiently to conventional therapies | Additional indication |
| VIEKIRAX | Ombitasvir hydrate/Paritaprevir hydrate/Ritonavir | Manufacturer and distributor: AbbVie GK | September 28, 2015 | $204/pill | Improvement of viremia in patients with chronic hepatitis C or compensated cirrhosis type C in serogroup 1 (genotype 1) | New approval |
| CIPROXAN | Ciprofloxacin | Manufacturer and distributor: Bayer Yakuhin | September 24, 2015 | $0.3 (100mg/pill)  $0.4 (200mg/pill)  $17 (200mg/100ml)  $21 (300mg/150ml)  $19 (400mg/200ml) | Treatment of sepsis, pneumonia, etc. | Additional indication |
| HARVONI | Ledipasvir acetonate/Sofosbuvir | Manufacturer and distributor: Gilead Sciences | July 3, 2015 | $509/pill | Improvement of viremia in patients with chronic hepatitis C or compensated cirrhosis type C in serogroup 1 (genotype 1) | New approval |
| DIFLUCAN | Fluconazole | Manufacturer and distributor: Pfizer | May 26, 2015 | $3 (50mg capsule)  $4 (100mg capsule)  $17 (0.1% 50ml bottle)  $22 (0.2% 50ml bottle)  $36 (0.2% 100ml bottle) | Treatment of vaginitis and vulvovaginitis caused by Candida | Additional indication |
| ALDREB | Colistin sodium methanesulfonate | Manufacturer and distributor: GlaxoSmithKline K. K | March 26, 2015 | $77 (150mg bottle) | Treatment of infections caused by colistin-sensitive Escherichia coli, Citrobacter, Klebsiella, Enterobacter, Pseudomonas aeruginosa and Acinetobacter | New approval |
| SYNFLORIX | Pneumococcal 10-valent conjugate vaccine adsorbed | Manufacturer: GlaxoSmithKline K. K  Distributor: Daiichi Sankyo | March 26, 2015 | NA | Prophylaxis of pneumonia and pneumococcal invasive diseases | New approval |
| KAKETSUKEN | Cell culture-derived influenza emulsion HA vaccines (prototype) | Manufacturer and distributor: The Chemo-Sero-Therapeutic Research Institute | March 26, 2015 | NA | Prevention of pandemic influenza | New approval |
| SOVALDI | Sofosbuvir | Manufacturer and distributor: Gilead Sciences | March 26, 2015 | $395 (400mg pill) | Improvement of viremia in patients with chronic hepatitis C or compensated cirrhosis type C in serogroup 2 (genotype 2) | New approval |
| COPEGUS | Ribavirin | Manufacturer and distributor: Chugai Pharmaceutical | March 26, 2015 | $6 (200mg/pill) | Improvement of viremia with the concomitant use of sofosbuvir in patients with chronic hepatitis C or compensated cirrhosis type C in serogroup 2 (genotype 2) | Additional indication |
| DAKLINZA | Daclatasvir hydrochloride | Manufacturer and distributor: Bristol Myers Squibb | March 20, 2015 | $74 (60mg/pill) | Improvement of viremia in patients with chronic hepatitis C or compensated cirrhosis type C in serogroup 1 (genotype 1) | Changed approval |
| TRIUMEQ | Dolutegravir sodium, Abacavir sulfate, Lamivudine | Manufacturer and distributor: ViiV Healthcare | March 16, 2015 | $64/pill | Treatment of HIV infection | New approval |
| VENOGLOBULIN | Polyethylene glycol treated human normal immunoglobulin | Manufacturer and distributor: Japan Blood Products Organization | February 2, 2015 | $347 (5g/100ml bottle)  $702 (10g/200ml bottle) | Prevention of acute otitis media, acute bronchitis, or pneumonia caused by Pneumococcus or Hemophilus influenzae in patients associated with a decrease in serum IgG2 levels | Additional indication |

*Price per drug was converted into US dollars using the 2019 average monthly exchange rates of ¥109.0 per $1.

Price per drug was used as of February 19, 2022.

Drug price for vaccines was not determined and was open priced in Japan. So, it was not available.

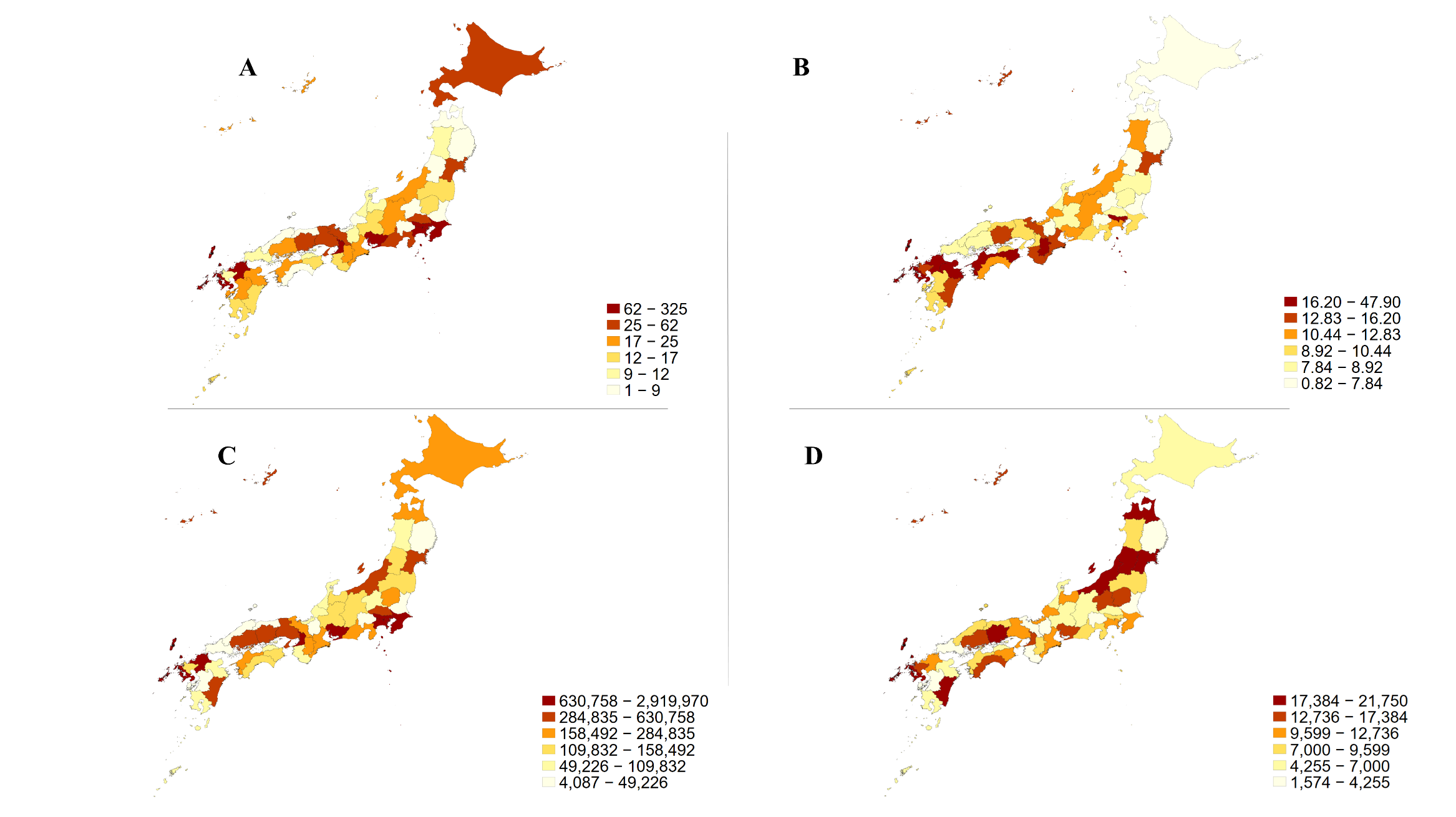

Supplemental Material 5. Geographical characteristics of payment distribution

5A: The number of infectious disease specialists in 2021; 5B: The number of infectious disease specialists per million population in 2021; 5C: total personal payment values to the infectious disease specialists from 2016 to 2019; 5D: average personal payment values per infectious disease specialist from 2016 to 2019
